# Evaluation of Artificial Intelligence (AI)-based *in silico* tools for variant classification in clinically actionable NSCLC variants

**DOI:** 10.1101/2024.04.12.24305738

**Authors:** Eugene Kim, Samantha E Duarte, Emma Yu, Ilene Hong, Grace Lee, Young Kwang Chae

**Affiliations:** Northwestern University Feinberg School of Medicine, Chicago, IL; Northwestern University, Evanston, IL; Department of Hematology and Oncology, Department of Medicine, Northwestern University Feinberg School of Medicine, Chicago, IL; Robert H. Lurie Comprehensive Cancer Center of Northwestern University, Chicago, IL; Department of Internal Medicine, Northwestern University Feinberg School of Medicine, Chicago, IL

## Abstract

**Introduction/Background:** Genetic variants beyond FDA-approved drug targets are often identified in non-small cell lung cancer (NSCLC) patients. Although the performances of *in silico* tools in predicting variant pathogenicity have been analyzed in previous studies, they have not been analyzed for actionable targets of FDA-approved therapies for NSCLC. The aim of this study is to compare the performance of commonly used *in silico* tools in classifying the pathogenicity of actionable variants in NSCLC.

**Materials and Methods:** We evaluated the performance of the following *in silico* tools: Polyphen-2 (HumDiv, HumVar), Align-GVGD, MutationTaster2021, CADD, CONDEL, and REVEL. A curated set of targetable NSCLC missense variants (n=236) was used. The overall accuracy, sensitivity, specificity, positive predictive value (PPV), negative predictive value (NPV), and Matthews correlation coefficient (MCC) of each *in silico* tool was determined.

**Results:** The most recently released MutationTaster2021 demonstrated the highest performance in terms of accuracy, specificity, PPV, and MCC, but was outperformed by CADD for both sensitivity and NPV. Although some tools demonstrated high sensitivities, all tools except MutationTaster2021 displayed markedly low overall specificities, as low as 23%.

**Conclusion:** The collective results indicate that the evaluated *in silico* tools can provide guidance in predicting the pathogenicity of NSCLC missense variants, but are not fully reliable. The tools analyzed in this study could be acceptable to rule out pathogenicity in variants given their higher sensitivities, but are limited when it comes to identifying pathogenicity in variants due to low specificities.

**Highlights:**

- MutationTaster2021 demonstrated the highest overall performance
- CADD demonstrated the highest sensitivity (99.19%) and NPV (96.55%).
- All tools but MutationTaster2021 demonstrated low specificities, as low as 23%
- An individual predictor outperformed 3 meta predictors
- Performance variability suggests caution when using *in silico* tools in the clinic

## Introduction

Next generation sequencing continues to play a pivotal role in routine practice in the era of precision medicine [1, 2]. As our ability to sequence the genome has advanced, genetic variants beyond FDA-approved drug targets and variants of uncertain significance (VUS) are often identified in patients [3]. To address the increasing complexity of interpreting genomic data, the use of methods such as molecular tumor boards (MTBs) involving an interdisciplinary review committee to guide personalized treatment decisions has evolved. However, given the limitations and discordance of MTBs, the need for better tools to translate immense amounts of genomic data into actionable clinical decisions remains a prevalent issue [4, 5]. As such, the American College of Medical Genetics and Genomics (ACMG) has published standards and guidelines for germline variant classification to provide a more standardized framework for variant interpretation and guide clinical decision making [6]. Even so, the lack of sufficient evidence such as segregation analysis and literature evidence of functional impact for rare variants remains a challenge to evaluating pathogenicity in the clinical setting. The accurate interpretation of VUS is especially crucial in precision oncology and plays a large role in clinical decision making for patients with variants beyond drug-approved targets [7]. The need for accurate methods to identify actionable driver mutations for precision therapy remains a prevalent issue in practical cancer management.

To address this challenge, numerous *in silico* variant classification prediction tools have been developed utilizing artificial intelligence (AI)-based algorithms that predict whether specific variants contribute to disease pathogenicity. Many of these tools aim to predict whether a non-synonymous single nucleotide variant in a coding region has a phenotypic effect in relation to pathogenicity, and can generally predict hundreds or more variants simultaneously. Among the variant classification tools available, there are widely different AI-based approaches. One approach relies on evolutionary principles, categorizing variants based on factors such as multiple sequence alignments, conservation of the amino acid through evolution, and high or low allele frequency. Tools that utilize such methods include DANN [8], PrimateAI [9], and PANTHER [10]. Another approach incorporates the effect of variants on aspects of protein function by affecting properties such as polarity, charge, genomic location in relation to functional regions, and 3-D structure. iStable [11], SNPs&GO [12], and Mutpred2 [13] are examples of protein structure/function-analyzing tools. Commonly used tools take a combined approach to variant classification, as seen in PolyPhen-2 [14], Align-GVGD [15], and MutationTaster2021 [16]. For prediction of nsSNPs, recent developments have begun to analyze changes in splice sites, chromatin effects, and patterns in regulatory motifs. DeepSEA [17], NetGene2 [18], and DanQ [19] are such examples. Meta-predictors such as Revel [20], BayesDel [21], Condel [22], and CADD [23] consolidate multiple classifiers and have been shown to perform better than individual *in silico* predictors alone [24, 25].

ACMG/AMP *Standards and Guidelines* indicate the cautious use of these publicly and commercially available *in silico* tools to predict the classification of rare variants. Predictions from all *in silico* programs must agree, otherwise the evidence should not be used as stated by the guidelines. Evidence from *in silico* tools are included as one of the criteria for variant classification, as there are 28 criteria that address evidence for variant classification as defined by the guidelines, each categorized by weight and type. The criteria divided by type are: population data, computational data, functional data, segregation data, *de novo* data, allelic data, other database, and other data. Variants are then classified into five tiers according to applied criteria: pathogenic (P), likely pathogenic (LP), variant of uncertain significance (VUS), likely benign (LB), and benign (B) [6].

Understanding the performance and reliability of i*n silico* tools is imperative as accuracy of computational methods have direct implications on clinical management involving patients with VUS. Although studies have shown variable results regarding the performance of *in silico* tools, recent studies have shown the need to further evaluate these tools in a clinical context [26, 27].

Although *in silico* variant prediction tools have been analyzed in previous studies, the performance of such predictors has not been analyzed for variants of actionable targets of NSCLC. The aim of this study is to evaluate the performance of four individual and three combined *in silico* classification tools for predicting the pathogenicity of 236 missense variants of clinically actionable genes associated with NSCLC.

## Materials and Methods

### Variant Selection

NSCLC genes of interest were included for evaluation based on indications as molecular targets in the National Comprehensive Cancer Network (NCCN) Guidelines for NSCLC [28]. Our initial dataset consisted of 296 variants of *BRAF, EGFR, ERBB2, KRAS, MET, ALK, ROS1, NTRK1, NTRK2,* and *NTRK3* genes. Pathogenic variants were curated based on annotations that specifically indicated pathogenicity for NSCLC in the NCCN Guidelines, OncoKB, and My Cancer Genome [28, 29, MCG; mycancergenome.org]. ClinVar was not used to select pathogenic variants due to the database’s low volume of pathogenic and likely pathogenic variants specifically for NSCLC. Benign NSCLC variants were curated from the dbSNP database with the inclusion criteria of a benign or likely benign ClinVar assertion. These sources were used to define a true classification for the variants in our dataset. A final dataset of 236 pathogenic (n=136) and benign (n=100) NSCLC variants was used for analysis.

### *In silico* Classification Tool Selection

Polyphen-2 (HumDiv, HumVar) [14], Align-GVGD [15], MutationTaster2021 [16], CADD [23], CONDEL [22], and REVEL [20] were the *in silico* classification tools evaluated in this study. *In silico* tools were largely selected based on inclusion in the ACMG/AMP *Standards and Guidelines* [6] and common use based on literature review. Polyphen-2, Align-GVGD, and MutationTaster2021 are individual tools for pathogenicity prediction, while CADD, CONDEL, and REVEL are meta-predictors that incorporate a combination of individual scores to classify variants. CADD combines 60 distinct annotations including Ensembl Variant Effect Predictor (VEP), phyloP, phastCons, GERP++, Grantham, SIFT and Polyphen-2. CONDEL outputs the weighted average of the scores of MutationAssessor and FATHMM. REVEL, which integrates scores from MutPred, FATHMM v2.3, VEST 3.0, PolyPhen-2, SIFT, PROVEAN, MutationAssessor, MutationTaster, LRT, GERP++, SiPhy, phyloP, and phastCons, was included in this study based on its superior performance in a recent study by Tian et al. [30]. Of note, MutationTaster2021, the latest update to MutationTaster, was implemented for this study due to its improved prediction model and cited improvements in accuracy [16].

### Parameter Setting

The default thresholds as suggested by the tools’ authors were used to classify variants. Pathogenicity for tools that gave a numerical score was defined as a score <0.05 for Polyphen-2, ≥C35 for Align-GVGD, >15 for CADD, and >0.05 for REVEL. The categorical classifications of Polyphen-2, MutationTaster2021 and CONDEL were used directly from their outputs.

Polyphen-2 classified variants as Probably Damaging, Possibly Damaging, and Benign. MutationTaster2021 classified variants as Deleterious, Deleterious (ClinVar), Benign, or Benign (auto). CONDEL classified variants as either Deleterious or Neutral.

### Evaluation of Performance

Each missense variant in the final dataset was classified using each *in silico* tool. True positive (TP) results refer to the correct prediction of pathogenicity as defined in our final dataset using the criteria mentioned above. True negative (TN) results refer to the correct prediction of benign variants as defined by the criteria mentioned above. The following measures were obtained for each tool: overall accuracy 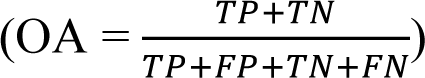, and sensitivity 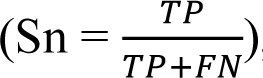, specificity (Sp = 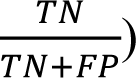, and Matthews correlation coefficient 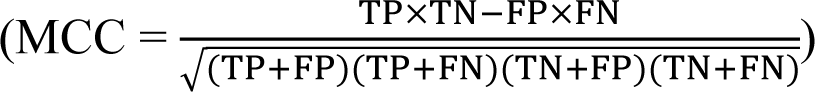 were calculated.

Sensitivity (true positive rate) was reflective of each *in silico* tool’s ability to identify true pathogenic variants, while specificity (true negative rate) demonstrated each tool’s ability to identify true benign variants. Results that yielded errors in calling predictions or had conflicting interpretations within the same tool were excluded from analysis and recorded as an error. MCC contributed a metric for balanced comparison as it accounts for the different sample sizes for each tool, as well as the imbalanced proportion of pathogenic to benign variants within our dataset. MCC values range from -1 (always false) to +1 (always correct), with a value of 0 indicating a completely random classification.

## Results

### Characteristics of selected variants

A summary of all variants evaluated in this study and source evidence can be found in Table S1. While our initial dataset consisted of 256 variants, the final dataset of 236 variants (136 pathogenic, 100 benign) was curated by removing variants with algorithmic interpretation errors or conflict. Errors included algorithms calling predictions for wrong amino acid changes despite correct inputs and conflicts within MutationTaster2021’s predictions based on transcript variants for the same gene. Notably, all 20 variants of *NTRK1*, *NTRK2*, and *NTRK3* were removed from analysis given the large proportion of variants with interpretation errors among all algorithms, including an inability to predict the specific variants desired. ClinVar provided source evidence for all benign variants. MyCancerGenome provided source evidence for all pathogenic variants of *BRAF, ERBB2,* and *ROS1*; it provided source evidence for the majority of pathogenic variants of *KRAS* (98.04%), *EGFR* (85.71%), and *ALK* (54.55%). *MET* variant pathogenicity was supported by OncoKB. All *ALK* variants included in this study were resistance mutations.

### Overall performance

To determine overall performance for each algorithm we analyzed accuracy, sensitivity, specificity, PPV, NPV, and MCC using all 236 variants included in our study. The results of this analysis can be found in Table 1 and Figure 1. MutationTaster2021 achieved the highest overall performance profile in terms of accuracy (92.76%), specificity (91.75%), PPV (93.55%), and MCC (0.85); it was outperformed by CADD for both sensitivity (99.19%) and NPV (96.55%). In contrast, Align-GVGD had the lowest overall performance profile in terms of accuracy, specificity, PPV, NPV, and MCC. Align-GVGD misclassified 112/236 variants (35 pathogenic, 77 benign). CONDEL achieved the lowest sensitivity. Pholyphen-2 (HumDiv), Polyphen-2 (HumVar), MutationTaster2021, CADD, and REVEL each achieved a sensitivity >90%.

**Table 1.** Overall and Gene-Specific Performance of *In Silico* Variant Classification Tools.

| Algorithm | Total variants (n) | Accuracy (%) | Sensitivity (%) | Specificity (%) | PPV (%) | NPV (%) |
| --- | --- | --- | --- | --- | --- | --- |
| **Overall performance** |  |  |  |  |  |  |
| PolyPhen-2 (HumDiv) | 235/236 | 80.00 | 94.12 | 60.61 | 76.65 | 88.24 |
| PolyPhen-2 (HumVar) | 235/236 | 81.28 | 89.71 | 69.70 | 80.26 | 83.13 |
| Align-GVGD | 236/236 | 52.54 | 74.26 | 23.00 | 56.74 | 39.66 |
| MutationTaster2021 | 221/236 | 92.76 | 93.55 | 91.75 | 93.55 | 91.75 |
| CONDEL | 223/236 | 71.75 | 72.36 | 71.00 | 75.42 | 67.62 |
| CADD | 222/236 | 67.57 | 99.19 | 28.28 | 63.21 | 96.55 |
| REVEL | 225/236 | 88.89 | 99.20 | 76.00 | 83.78 | 98.70 |
| **BRAF** |  |  |  |  |  |  |
| PolyPhen-2 (HumDiv) | 31/32 | 83.87 | 94.12 | 71.43 | 80.00 | 90.91 |
| PolyPhen-2 (HumVar) | 31/32 | 80.65 | 76.47 | 85.71 | 86.67 | 75.00 |
| Align-GVGD | 32/32 | 56.25 | 82.35 | 26.67 | 56.00 | 57.14 |
| MutationTaster2021 | 28/32 | 89.29 | 92.86 | 85.71 | 86.67 | 92.31 |
| CONDEL | 28/32 | 82.14 | 92.31 | 73.33 | 75.00 | 91.67 |
| CADD | 28/32 | 53.57 | 100 | 7.14 | 51.85 | 100 |
| REVEL | 29/32 | 96.55 | 100 | 93.33 | 93.33 | 100 |
| **EGFR** |  |  |  |  |  |  |
| PolyPhen-2 (HumDiv) | 36/36 | 94.44 | 96.43 | 87.50 | 96.43 | 87.50 |
| PolyPhen-2 (HumVar) | 36/36 | 94.44 | 96.43 | 87.50 | 96.43 | 87.50 |
| Align-GVGD | 36/36 | 66.67 | 75.00 | 37.50 | 80.77 | 30.00 |
| MutationTaster2021 | 33/36 | 90.91 | 88.00 | 100 | 100 | 72.73 |
| CONDEL | 33/36 | 54.55 | 48.00 | 75.00 | 85.71 | 31.58 |
| CADD | 33/36 | 93.94 | 100 | 75.00 | 92.59 | 100 |
| REVEL | 33/36 | 93.94 | 100 | 75.00 | 92.59 | 100 |
| **ERBB2 (HER2)** |  |  |  |  |  |  |
| PolyPhen-2 (HumDiv) | 32/32 | 87.50 | 92.00 | 71.43 | 92.00 | 71.43 |
| PolyPhen-2 (HumVar) | 32/32 | 81.25 | 84.00 | 71.43 | 91.30 | 55.56 |
| Align-GVGD | 32/32 | 53.13 | 64.00 | 14.29 | 72.73 | 10.00 |
| MutationTaster2021 | 30/32 | 90.00 | 91.30 | 85.71 | 95.45 | 75.00 |
| CONDEL | 30/32 | 43.33 | 43.48 | 42.86 | 71.43 | 18.75 |
| CADD | 29/32 | 79.31 | 95.45 | 28.57 | 80.77 | 66.67 |
| REVEL | 30/32 | 80.00 | 95.65 | 28.57 | 81.48 | 66.67 |
| **KRAS** |  |  |  |  |  |  |
| PolyPhen-2 (HumDiv) | 53/53 | 90.57 | 90.20 | 100 | 100 | 28.57 |
| PolyPhen-2 (HumVar) | 53/53 | 90.57 | 90.20 | 100 | 100 | 28.57 |
| Align-GVGD | 53/53 | 75.47 | 78.43 | 0 | 95.24 | 0 |
| MutationTaster2021 | 49/53 | 93.88 | 95.74 | 50.00 | 97.83 | 33.33 |
| CONDEL | 49/53 | 95.92 | 95.74 | 100 | 100 | 50.00 |
| CADD | 49/53 | 95.92 | 100 | 0 | 95.92 | undefined |
| REVEL | 50/53 | 96.00 | 100 | 0 | 96.00 | undefined |
| **MET** |  |  |  |  |  |  |
| PolyPhen-2 (HumDiv) | 16/16 | 68.75 | 100 | 64.29 | 28.57 | 100 |
| PolyPhen-2 (HumVar) | 16/16 | 81.25 | 100 | 64.29 | 40.00 | 100 |
| Align-GVGD | 16/16 | 25.00 | 50.00 | 21.43 | 8.33 | 75.00 |
| MutationTaster2021 | 16/16 | 87.50 | 100 | 85.71 | 50.00 | 100 |
| CONDEL | 16/16 | 75.00 | 100 | 71.43 | 33.33 | 100 |
| CADD | 16/16 | 37.50 | 100 | 28.57 | 16.67 | 100 |
| REVEL | 16/16 | 37.50 | 100 | 28.57 | 16.67 | 100 |
| **ALK** |  |  |  |  |  |  |
| PolyPhen-2 (HumDiv) | 45/45 | 73.33 | 100 | 64.71 | 47.83 | 100 |
| PolyPhen-2 (HumVar) | 45/45 | 77.78 | 100 | 70.59 | 52.38 | 100 |
| Align-GVGD | 45/45 | 35.56 | 72.73 | 23.53 | 23.53 | 72.73 |
| MutationTaster2021 | 43/45 | 100 | 100 | 100 | 100 | 100 |
| CONDEL | 45/45 | 73.33 | 63.64 | 76.47 | 46.67 | 86.67 |
| CADD | 45/45 | 51.11 | 100 | 35.29 | 33.33 | 100 |
| REVEL | 45/45 | 97.78 | 100 | 97.06 | 91.67 | 100 |
| **ROS1** |  |  |  |  |  |  |
| PolyPhen-2 (HumDiv) | 22/22 | 31.82 | 100 | 25.00 | 11.76 | 100 |
| PolyPhen-2 (HumVar) | 22/22 | 45.45 | 100 | 40.00 | 14.29 | 100 |
| Align-GVGD | 22/22 | 22.73 | 50.00 | 20.00 | 5.88 | 80.00 |
| MutationTaster2021 | 22/22 | 90.91 | 100 | 90.00 | 50.00 | 100 |
| CONDEL | 22/22 | 63.64 | 50.00 | 65.00 | 12.50 | 92.86 |
| CADD | 22/22 | 22.73 | 100 | 15.00 | 10.53 | 100 |
| REVEL | 22/22 | 86.36 | 100 | 85.00 | 40.00 | 100 |

**Figure 1.**
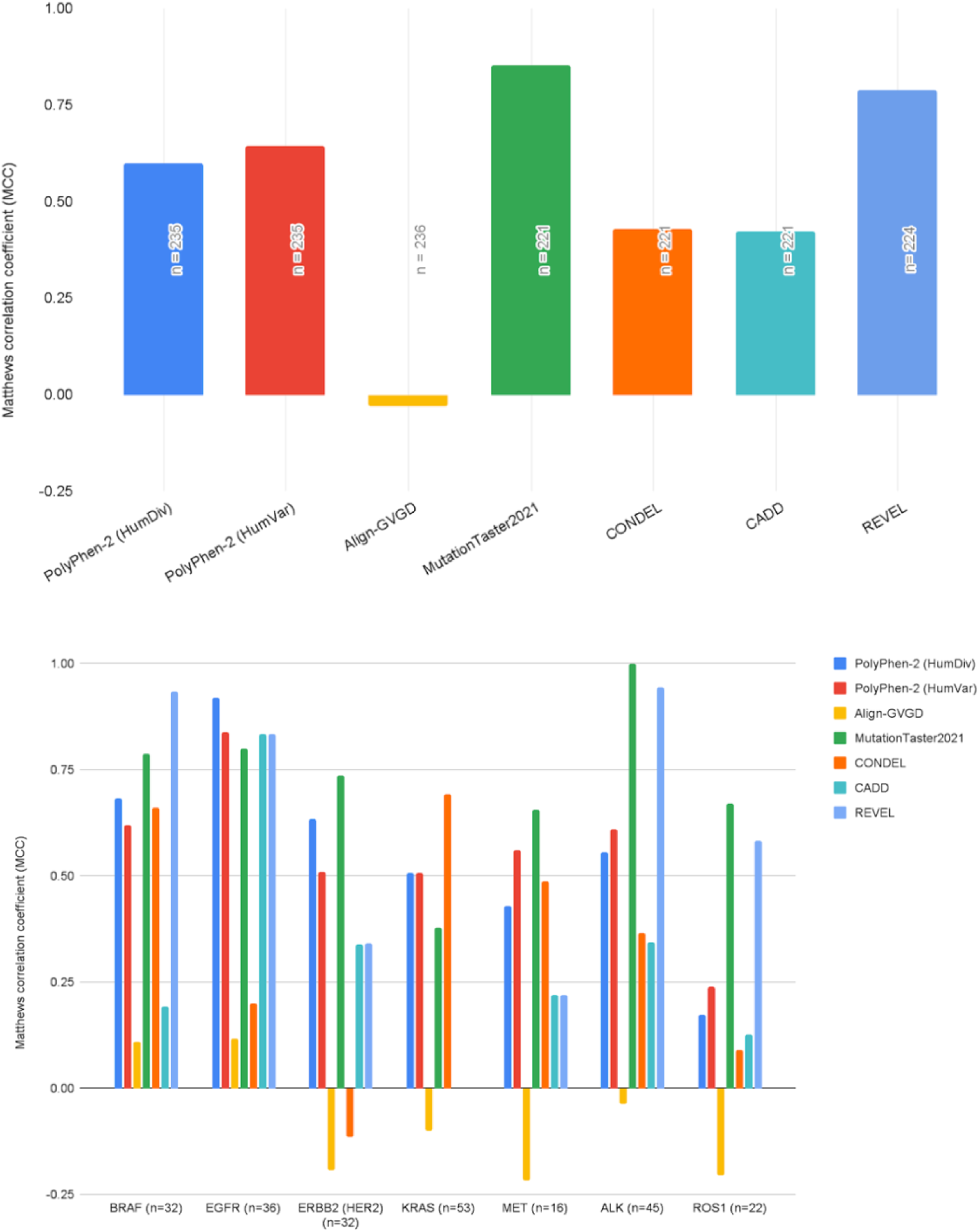
Overall (A) and Gene-Specific (B) Performance of *In Silico* Variant Classification Tools by Matthew’s Correlation Coefficient. Performance of individual (Polyphen-2, Align-GVGD, MutationTaster2021) and combined (CADD, CONDEL, REVEL) variant classification tools was evaluated using a dataset of 236 pathogenic (n=136) and benign (n=100) variants of clinically actionable genes associated with NSCLC (*BRAF, EGFR, ERBB2, KRAS, MET, ALK, ROS1, NTRK1, NTRK2, NTRK3*). Results that yielded errors in calling predictions or had conflicting interpretations within the same tool were excluded from analysis as represented by total variants in Table 1. MCC values range from -1 (always false) to +1 (always correct), with a value of 0 indicating a completely random classification. A complete list of variants included in the study can be found in Supplementary Table S1.

Furthermore, CADD achieved a sensitivity of 99.19%. In terms of specificity, Align-GVGD and CADD achieved scores <30%. Similar MCC scores were observed between PolyPhen-2 (HumDiv) and PolyPhen-2 (HumVar) (0.60 and 0.61, respectively), as well as CADD and CONDEL (0.41 and 0.43, respectively). Align-GVGD was the sole algorithm to achieve an MCC score <0 (−0.03). Concordance among all tools was observed for 32.56% of variants.

### Performance of all tools based on genes

We also evaluated the performance of each algorithm by gene for accuracy, sensitivity, specificity, PPV, NPV, and MCC. The results of this analysis can be found in Table 1 and Figure 1.

Errors in *BRAF* variant interpretation were observed among all algorithms; the fewest errors were observed with PolyPhen-2 (HumDiv) and PolyPhen-2 (HumVar) (1/32 variants excluded), while Align-GVGD, MutationTaster2021, CONDEL, and CADD had the most errors (4/32 variants excluded). REVEL achieved the highest performance for *BRAF* variants for all performance measurements. All algorithms achieved a sensitivity of >80% with CADD and REVEL achieving sensitivity of 100%. Specificity <30% were observed with Align-GVGD and CADD.

Errors in *EGFR* variant interpretation were observed with MutationTaster2021, CONDEL, CADD, and REVEL (3/36 variants excluded). PolyPhen-2 (HumDiv), PolyPhen-2 (HumVar), CADD, and REVEL achieved sensitivity >90%. For specificity, all algorithms achieved values ≥75% except Align-GVGD. MCC scores were ≥0.80 for PolyPhen-2 (HumDiv), PolyPhen-2 (HumVar), MutationTaster 2021, CADD, and REVEL. Notably, MCC scores for Align-GVGD and CONDEL were ≤0.20.

Errors in *ERBB2* variant interpretation were observed with MutationTaster2021, CONDEL, Revel (2/32 variants excluded), while CADD had the most errors (3/32 variants excluded). PolyPhen-2 (HumDiv), MutationTaster2021, REVEL, and CADD achieved sensitivities ≥90%.

Specificity <30% were observed with Align-GVGD, CADD and REVEL. Of note, MCC scores <0 were achieved by Align-GVGD and CONDEL (−0.19 and -0.12, respectively).

Errors in KRAS variant interpretation were observed with CADD (3/53 variants excluded), while the most errors were seen with MutationTaster2021, CONDEL, and CADD (4/53 variants excluded). All algorithms achieved an accuracy and sensitivity of ≥90% except Align-GVGD. A specificity of 100% was observed for Polyphen-2 (HumDiv), PolyPhen-2 (HumVar), and CONDEL. In contrast, specificity of 0% was observed with Align-GVGD, CADD, and REVEL. Of note, MCC scores <0 were achieved by Align-GVGD (−0.10), while MCC scores for CADD and REVEL were undefined.

Align-GVGD achieved the lowest performance for MET variants for all performance measures. All other algorithms achieved a sensitivity of 100%. Specificity <30% were observed with Align-GVGD, CADD and REVEL.

Errors in ALK variant interpretation were observed with MutationTaster2021 (2/45 variants excluded). MutationTaster2021 and REVEL showed high performance for ALK variants for all performance measures. Sensitivity of 100% was also achieved by Polyphen-2 (HumDiv), PolyPhen-2 (HumVar), and CADD. Align-GVGD achieved the lowest performance for all performance measures except sensitivity.

MutationTaster2021 achieved the highest overall performance for ROS1 variants for all performance measures. Align-GVGD had the lowest performance for all performance measures except specificity. Sensitivity of 100% was achieved by all algorithms, except Align-GVGD and CONDEL. Specificity <30% were observed with Polyphen-2 (HumDiv), Align-GVGD, and CADD.

Errors in variant-calling were observed among all algorithms for most *NTRK1*, *NTRK2*, and *NTRK3* variants. All algorithms failed to interpret >50% of *NTRK1* variants, ranging from 55.56% of target *NTRK1* variants unable to be predicted (MutationTaster2021, CADD) to 77.78% of *NTRK1* target variants (Align-GVGD). All successful variant predictions resulted in false postives for PolyPhen-2 (HumDiv), PolyPhen-2 (HumVar), Align-GVGD, and CADD. MutationTaster2021, CONDEL, CADD, and REVEL failed to interpret *NTRK2* and *NTRK3* variants.

## Discussion

In this study, we evaluated the performance of four individual and three combined *in silico* classification tools for predicting the pathogenicity of 236 missense variants of clinically actionable genes associated with NSCLC. By specifically targeting NSCLC variants in our study, this study provides a practical and curated analysis on the performance of commonly used *in silico* tools within the context of clinical precision medicine. Our findings suggest that an individual predictor outperformed 3 meta-predictors, which deviates from the results of other studies [24, 25]. MutationTaster2021 outperformed CONDEL, CADD, and REVEL for overall performance and 4 of 7 genes (*ERBB2, MET, ALK, ROS1*) (Table 1). In contrast, Align-GVGD demonstrated poorer performance overall and for 6 of 7 genes (*EGFR, ERBB2, KRAS, MET, ALK, ROS1*). This study deviates from prior reports of Align-GVGD outperforming other variant classification tools in terms of overall specificity, accuracy, and MCC [26, 27]. However, other studies report low performance of Align-GVGD compared to other predictors [31]. Of interest, MutationTaster2021 released on 07/02/2021 is a newly updated tool replacing the previous MutationTaster2, stated to have higher accuracy than its predecessor by its authors [16]. As disclosed on their website, Align-GVGD was last updated on 09/08/2014. The results displayed by the newest and oldest tools in our study may be demonstrative of both the speed of growth in the field of precision genomics, as well as the need for tools to keep up-to-date as our knowledge of pathogenic variants continues to grow.

The results of our study align with similar studies which have also shown shortcomings of *in silico* prediction tools for genetic testing. Broadly, even if a tool demonstrated high sensitivity, all tools except MutationTaster2021 displayed markedly low overall specificities, as low as 23.00%. This result is consistent with two other recent studies analyzing the performance of similar tools in a clinical context [26, 27]. According to our results, interpretation of gene variants related to *BRAF, EGFR, ERBB2, KRAS, MET, ALK,* and *ROS1* using our highest performing tool (MutationTaster2021) would result in 8 false negative results out of 136 patients with a true pathogenic mutation related to NSCLC, and 8 false positive results in 100 patients with true benign mutations. When translated to real clinical decision making, the significance of a false negative prediction can result in a patient missing out on a potentially effective treatment option. On the other hand, a false positive can result in a sub-optimal treatment for a patient which has implications for worse outcomes, increased cost of care, as well as increased chance of toxicity. Additionally, we observed significant variability in tool performance among variants of different genes which demonstrates the lack of a single tool that performed well among all testing genes. Evaluation of *in silico* variant prediction tools is clinically relevant given a number of biotechnology companies involved in commercial gene profiling employ tools such as MutationTaster2021, Polyphen, and Align-GVGD to provide physicians with variant predictions that help guide targeted therapy.

During our quality control check, we found 43 results with pathogenicity predictions that differed from the initial results. The majority of these inconsistencies did not affect our final pathogenicity prediction. For example, Polyphen-2 (HumDiv) read *BRAF* V600E as “probably damaging” during the initial run, but “possibly damaging” during the QC check. However, we did identify 14 results leading to a change in pathogenicity prediction (9 pathogenic → benign; 5 benign → pathogenic). The findings of this quality control check speaks to the ever-increasing amount of evidence that is utilized by these tools and the speed at which tools are improving with ongoing research. Apart from major updates by developers, lines of evidence are constantly evolving to the point that predictions can also evolve to reflect these advancements. Even without official updates released by the developers, *in silico* tools are constantly improving at a pace that seems encouraging for the future of clinical care.

Performance of *in silico* classification tools has clinical relevance as many biotech companies utilize both publicly available and proprietary prediction tools in evaluating patient samples. We encountered a number of issues during our analysis that could potentially limit the use of these platforms in such settings. We identified multiple variants that could not be interpreted by certain algorithms leading to exclusion from our analysis. Errors in variant interpretation may happen for a variety of reasons. Tools are often limited in the types of mutations they can evaluate.

Additionally, variant predictors often require external data which requires gene identifiers to be linked across databases. We removed *NTRK1*, *NTRK2*, and *NTRK3* from our final analysis due to the large proportion of variants with errors in variant interpretation. Our study highlights potential gaps in variant prediction tools, which may limit usefulness in evaluating a large proportion of variants of well-described, clinically relevant genes.

Our study was limited by the type of variants included, namely missense mutations. A number of well-studied, clinically actionable variants of genes related to NSCLC with approved targeted therapies were excluded from the study (e.g. *EGFR* exon 19 indel, *MET* exon 14 skipping mutations, *EML4-ALK* fusion, *ROS1* rearrangements). Although some *in silico* tools are capable of analyzing mutations beyond just missense mutations (e.g. DANN and DeepSEA), there remains a need for multiple *in silico* variant classification tools that fulfill varying roles in assigning pathogenicity to variants across different mutation classes.

Regarding future studies, the utility of applying gene-level thresholds for predicting pathogenicity in a practical clinical context may be a topic of interest. Previous studies have shown that using a global threshold across genes may lead to increases in false positive predictions compared to applying gene-level thresholds for assessing whether a VUS is benign or deleterious [32, 33]. In real clinical context, applying this level of precision may not be practical as gene-specific thresholds are not always readily available. However, if the utitlity of this approach is further explored, there may be improvements to be made in the application of *in silico* tools in precision medicine.

## Conclusion

To our knowledge, this is the first study to evaluate and compare the performance of AI-based *in silico* variant classification tools in predicting pathogenicity of clinically actionable missense variants associated with NSCLC. We observed significant variability in performance among all 6 tools, as well as internal performance variability among variants of different genes. Interestingly, we found MutationTaster2021 outperformed 3 meta predictors. This study has clinical implications as the decision to treat patients with NSCLC using immunotherapy or targeted therapy is often contingent upon determination of variant pathogenicity. Our results show that in silico variant classification tools may be useful in ruling out, but not ruling in, pathogenicity for VUS. *In silico* classification tools may provide guidance on clinical decisions for patients with VUS, but should be used with caution given the significant variability in pathogenicity predictions.

## Data Availability

All data produced in the present study are available upon reasonable request to the authors.

## Acknowledgements

Not applicable.

## Data Availability Statement

Data will be made available on request.

