## Supplementary Table S1 for "Evaluation of Artificial Intelligence (AI)-based *in silico* tools for variant classification in clinically actionable NSCLC variants"

### Supplementary Tables & Figures

**Table S1. NSCLC Missense Variants, Pathogenicity, and Source Evidence.**
NSCLC missense variants were included for evaluation based on indications as molecular targets in NCCN Guidelines for NSCLC (BRAF, EGFR, ERBB2, KRAS, MET, ALK, ROS1, NTRK1, NTRK2, and NTRK3). Benign NSCLC variants in our dataset were curated from the dbSNP database with the inclusion criteria of a benign or likely benign ClinVar assertion. Pathogenic variants were curated based on annotations in NCCN Guidelines 6.2021, OncoKB, and My Cancer Genome indicated specifically for NSCLC. “1” denotes pathogenic and “0” denotes benign based on source evidence. “*” denotes genes excluded from the final analysis.

| | Pathogenic (1) Benign (0) | Source evidence |
|---|---|---|
| **BRAF** | | |
| V600E | 1 | NCCN Guidelines |
| V600D | 1 | MyCancerGenome |
| V600K | 1 | MyCancerGenome |
| V600M | 1 | MyCancerGenome |
| V600R | 1 | MyCancerGenome |
| V600G | 1 | MyCancerGenome |
| Y472C | 1 | MyCancerGenome |
| K601E | 1 | MyCancerGenome |
| L597Q | 1 | MyCancerGenome |
| D594G | 1 | MyCancerGenome |
| L597R | 1 | MyCancerGenome |
| L597S | 1 | MyCancerGenome |
| L597V | 1 | MyCancerGenome |
| G469A | 1 | MyCancerGenome |
| G469V | 1 | MyCancerGenome |
| D594N | 1 | MyCancerGenome |
| G466V | 1 | MyCancerGenome |
| E26D | 0 | ClinVar |
| D22N | 0 | ClinVar |
| E26K | 0 | ClinVar |
| S147G | 0 | ClinVar |
| A31G | 0 | ClinVar |
| S323L | 0 | ClinVar |
| R384G | 0 | ClinVar |

| | | |
|---|---|---|
| E24D | 0 | ClinVar |
| P25S | 0 | ClinVar |
| I326V | 0 | ClinVar |
| R477Q | 0 | ClinVar |
| T332I | 0 | ClinVar |
| D742N | 0 | ClinVar |
| G30D | 0 | ClinVar |
| A34V | 0 | ClinVar |
| **EGFR** | | |
| L858R | 1 | NCCN Guidelines |
| L861Q | 1 | NCCN Guidelines |
| S768I | 1 | NCCN Guidelines |
| T790M | 1 | NCCN Guidelines |
| D761Y | 1 | OncoKB |
| L747P | 1 | OncoKB |
| C797S | 1 | OncoKB |
| C797G | 1 | OncoKB |
| G719A | 1 | Mycancergenome |
| G719C | 1 | Mycancergenome |
| G719S | 1 | Mycancergenome |
| G719D | 1 | Mycancergenome |
| A289T | 1 | Mycancergenome |
| E709K | 1 | Mycancergenome |
| L861R | 1 | Mycancergenome |
| V769M | 1 | Mycancergenome |
| K860I | 1 | Mycancergenome |
| L833V | 1 | Mycancergenome |
| V769L | 1 | Mycancergenome |
| V834L | 1 | Mycancergenome |
| V843I | 1 | Mycancergenome |
| A859S | 1 | Mycancergenome |
| A871E | 1 | Mycancergenome |
| G719E | 1 | Mycancergenome |
| H835L | 1 | Mycancergenome |
| H870R | 1 | Mycancergenome |
| L833F | 1 | Mycancergenome |
| L861G | 1 | Mycancergenome |
| R521K | 0 | ClinVar |
| H988P | 0 | ClinVar |

| | | |
|---|---|---|
| S1162N | 0 | ClinVar |
| A1048V | 0 | ClinVar |
| V904I | 0 | ClinVar |
| L184P | 0 | ClinVar |
| I966V | 0 | ClinVar |
| I491T | 0 | ClinVar |
| **ERBB2** | | |
| V842I | 1 | Mycancergenome |
| D769Y | 1 | Mycancergenome |
| I767M | 1 | Mycancergenome |
| S310F | 1 | Mycancergenome |
| V777L | 1 | Mycancergenome |
| L755S | 1 | Mycancergenome |
| D769H | 1 | Mycancergenome |
| S310Y | 1 | Mycancergenome |
| V659E | 1 | Mycancergenome |
| L869R | 1 | Mycancergenome |
| R678Q | 1 | Mycancergenome |
| V697L | 1 | Mycancergenome |
| V777M | 1 | Mycancergenome |
| T862A | 1 | Mycancergenome |
| T733I | 1 | Mycancergenome |
| D769N | 1 | Mycancergenome |
| L755P | 1 | Mycancergenome |
| I655V | 1 | Mycancergenome |
| L786V | 1 | Mycancergenome |
| T862I | 1 | Mycancergenome |
| V773M | 1 | Mycancergenome |
| G776V | 1 | Mycancergenome |
| G776C | 1 | Mycancergenome |
| L755A | 1 | Mycancergenome |
| R868W | 1 | Mycancergenome |
| I654V | 0 | ClinVar |
| W452C | 0 | ClinVar |
| A1216D | 0 | ClinVar |
| E79A | 0 | ClinVar |
| A386D | 0 | ClinVar |
| G815R | 0 | ClinVar |
| A1039T | 0 | ClinVar |

| **KRAS** | | |
|---|---|---|
| G12C | 1 | OncoKB |
| G12V | 1 | Mycancergenome |
| G13C | 1 | Mycancergenome |
| G13D | 1 | Mycancergenome |
| G12A | 1 | Mycancergenome |
| G12D | 1 | Mycancergenome |
| G12R | 1 | Mycancergenome |
| Q61H | 1 | Mycancergenome |
| Q61K | 1 | Mycancergenome |
| Q61L | 1 | Mycancergenome |
| Q61R | 1 | Mycancergenome |
| G12S | 1 | Mycancergenome |
| G13R | 1 | Mycancergenome |
| G13A | 1 | Mycancergenome |
| G13S | 1 | Mycancergenome |
| G13V | 1 | Mycancergenome |
| Q61P | 1 | Mycancergenome |
| A146T | 1 | Mycancergenome |
| G12F | 1 | Mycancergenome |
| G12L | 1 | Mycancergenome |
| G13E | 1 | Mycancergenome |
| A146P | 1 | Mycancergenome |
| A146V | 1 | Mycancergenome |
| K117N | 1 | Mycancergenome |
| A59T | 1 | Mycancergenome |
| A59E | 1 | Mycancergenome |
| A59G | 1 | Mycancergenome |
| Q61E | 1 | Mycancergenome |
| A59P | 1 | Mycancergenome |
| A59S | 1 | Mycancergenome |
| A59V | 1 | Mycancergenome |
| A146S | 1 | Mycancergenome |
| D33E | 1 | Mycancergenome |
| L19F | 1 | Mycancergenome |
| Q22K | 1 | Mycancergenome |
| T58I | 1 | Mycancergenome |
| V14I | 1 | Mycancergenome |
| D119N | 1 | Mycancergenome |

| | | |
|---|---|---|
| F156L | 1 | Mycancergenome |
| F28L | 1 | Mycancergenome |
| G60R | 1 | Mycancergenome |
| K147E | 1 | Mycancergenome |
| K5N | 1 | Mycancergenome |
| N116S | 1 | Mycancergenome |
| P34L | 1 | Mycancergenome |
| P34R | 1 | Mycancergenome |
| Q22E | 1 | Mycancergenome |
| Q22R | 1 | Mycancergenome |
| T74P | 1 | Mycancergenome |
| V14L | 1 | Mycancergenome |
| Y71H | 1 | Mycancergenome |
| G179S | 0 | ClinVar |
| Y166N | 0 | ClinVar |
| **MET** | | |
| D1228N | 1 | OncoKB |
| Y1230H | 1 | OncoKB |
| N375S | 0 | ClinVar |
| A1363T | 0 | ClinVar |
| E168D | 0 | ClinVar |
| M362T | 0 | ClinVar |
| A48V | 0 | ClinVar |
| H888Y | 0 | ClinVar |
| R1166Q | 0 | ClinVar |
| S572N | 0 | ClinVar |
| T733I | 0 | ClinVar |
| H1174R | 0 | ClinVar |
| N704D | 0 | ClinVar |
| S1015A | 0 | ClinVar |
| K632N | 0 | ClinVar |
| D543N | 0 | ClinVar |
| **ALK** | | |
| L1196M | 1 | OncoKB |
| I1171N | 1 | OncoKB |
| G1202R | 1 | OncoKB |
| C1156Y | 1 | OncoKB |
| G1269A | 1 | OncoKB |
| F1174L | 1 | Mycancergenome |

| | | |
|---|---|---|
| S1206Y | 1 | Mycancergenome |
| L1152R | 1 | Mycancergenome |
| F1174C | 1 | Mycancergenome |
| F1174V | 1 | Mycancergenome |
| L1198F | 1 | Mycancergenome |
| I1461V | 0 | ClinVar |
| K1491R | 0 | ClinVar |
| D1529E | 0 | ClinVar |
| P1599H | 0 | ClinVar |
| T1012M | 0 | ClinVar |
| V476A | 0 | ClinVar |
| T680I | 0 | ClinVar |
| V163L | 0 | ClinVar |
| E296K | 0 | ClinVar |
| E1419K | 0 | ClinVar |
| V198M | 0 | ClinVar |
| S426T | 0 | ClinVar |
| T648I | 0 | ClinVar |
| R259H | 0 | ClinVar |
| K1525E | 0 | ClinVar |
| P1027L | 0 | ClinVar |
| M1478T | 0 | ClinVar |
| P671S | 0 | ClinVar |
| R412C | 0 | ClinVar |
| A371T | 0 | ClinVar |
| R311H | 0 | ClinVar |
| G1494R | 0 | ClinVar |
| R510W | 0 | ClinVar |
| P36S | 0 | ClinVar |
| A1047T | 0 | ClinVar |
| S737L | 0 | ClinVar |
| E405D | 0 | ClinVar |
| P40S | 0 | ClinVar |
| N709K | 0 | ClinVar |
| S47L | 0 | ClinVar |
| P1029A | 0 | ClinVar |
| L1033H | 0 | ClinVar |
| V126A | 0 | ClinVar |
| L550F | 0 | ClinVar |

| | | |
|---|---|---|
| **ROS1** | | |
| D2033N | 1 | Mycancergenome |
| G2032R | 1 | Mycancergenome |
| E1902K | 0 | ClinVar |
| D1776H | 0 | ClinVar |
| S653F | 0 | ClinVar |
| N790S | 0 | ClinVar |
| R1506G | 0 | ClinVar |
| K2328R | 0 | ClinVar |
| P224S | 0 | ClinVar |
| K461E | 0 | ClinVar |
| N1095I | 0 | ClinVar |
| F942L | 0 | ClinVar |
| M792I | 0 | ClinVar |
| L760I | 0 | ClinVar |
| K1026M | 0 | ClinVar |
| K1890E | 0 | ClinVar |
| G1027D | 0 | ClinVar |
| D618G | 0 | ClinVar |
| P1659T | 0 | ClinVar |
| R2126Q | 0 | ClinVar |
| T804N | 0 | ClinVar |
| W142R | 0 | ClinVar |
| **NTRK1*** | | |
| H568Y | 0 | ClinVar |
| G577V | 0 | ClinVar |
| R744Q | 0 | ClinVar |
| R744P | 0 | ClinVar |
| M530T | 0 | ClinVar |
| T207M | 0 | ClinVar |
| R554C | 0 | ClinVar |
| P928L | 0 | ClinVar |
| R408Q | 0 | ClinVar |
| C246R | 0 | ClinVar |
| A722T | 0 | ClinVar |
| R55H | 0 | ClinVar |
| E245A | 0 | ClinVar |
| S160R | 0 | ClinVar |
| S174L | 0 | ClinVar |

| | | |
|---|---|---|
| R279C | 0 | ClinVar |
| R472W | 0 | ClinVar |
| T669S | 0 | ClinVar |
| **NTRK2*** | | |
| L140F | 0 | ClinVar |
| **NTRK3*** | | |
| V21F | 0 | ClinVar |
